# Application of bidirectional Mendelian randomization to assess the relationship between the gut microbiome and esophageal cancer

**DOI:** 10.1101/2025.02.06.25321825

**Authors:** James Chataway, Georgina Hamilton, Charlie Hatcher, Kaitlin H. Wade

## Abstract

**Background:** Esophageal cancer is the seventh most common cancer worldwide and typically carries a poor prognosis. Whilst smoking, alcohol and obesity are established risk factors, they do not fully account for disease variability and, increasingly, the gut microbiome has been implicated as a possible novel risk factor in observational studies. Mendelian randomization (MR), a technique that uses genetic variants as proxies for exposures to improve causal inference, is more robust to reverse causality and confounding, which typically bias observational studies.

**Methods:** We used summary statistics from large genome-wide association studies (GWASs) of both the gut microbiome and esophageal cancer phenotypes to conduct two-sample bidirectional MR analyses to investigate the causal relationship between 14 microbial traits and three esophageal cancer phenotypes: esophageal adenocarcinoma (EA), Barrett’s esophagus (BE) and both EA and BE as a combined phenotype (BE/EA). Where MR analyses provided evidence of causality between these phenotypes, several sensitivity analyses were conducted to interrogate its validity of MR assumptions.

**Results:** When assessing the causal role of the gut microbiome on esophageal cancer, there was little evidence that any microbial trait had a causal effect on any of the three esophageal cancer traits. In the reverse direction, MR analyses provided evidence that EA had a causal effect on two microbial traits. Specifically, an approximate doubling of the genetic liability to EA increased the odds of presence (vs. absence) of an unclassified group of bacteria within the *Firmicutes* phylum (odds ratio (OR): 1.66; 95% CI: 1.02, 2.70) and decreased the relative abundance of bacteria within the *Butyricicoccus* genus by 0.23 standard deviations (95% CI: 0.07, 0.40). However, importantly, sensitivity analyses showed that these observed effects were likely biased by horizontal pleiotropy and, thus, results should be interpreted with caution.

**Conclusions:** Although initial analyses provided evidence of EA influencing two microbial traits, further sensitivity analyses indicated that these results were likely biased and unlikely to reflect causality. This highlights the importance of using robust MR methodology with appropriate sensitivity analyses, particularly in the setting of microbial traits, where host genetic effects are poorly understood and likely to be complex.

## BACKGROUND

Esophageal cancer is the seventh most prevalent cancer and the sixth leading cause of cancer related death worldwide.(1,2) It is split histologically into two subtypes: esophageal squamous cell carcinoma (ESCC), commonly linked to alcohol consumption and smoking, and esophageal adenocarcinoma (EA), linked to gastro-esophageal reflux disease (GERD) and increased body weight.(3,4) Although ESCC accounts for the majority of esophageal cancer cases worldwide, EA is rapidly on the rise in Europe and the Western World(5–7), its incidence now surpassing ESCC in these populations.(8) This, accompanied with a typically poor prognosis and alarming mortality rates, means EA is fast becoming a major health crisis.(6,7) As a result, most recent studies of esophageal cancer involving European populations, including the largest genome-wide association study, focus on the EA histological subtype.(4,9) The histological pre-cursor to EA, Barrett’s esophagus (BE), is characterised by the transformation of esophageal squamous cells into metaplastic intestinal columnar epithelium.(10,11) However, less than 1% of cases of BE progress to EA (12,13), and current research fails to account for this gap in cases of EA. Therefore, identifying undiscovered novel risk factors and biological markers of EA will be key in reducing the global disease burden.

The gut microbiome and its influence on cancers of the gastrointestinal tract is mounting significant attention.(14,15) Several studies identified distinct differences in microbial species inhabiting the esophagus in individuals with and without BE and esophageal cancer (16,17), which could contribute to oncogenic and inflammatory processes.(18) Despite this growing body of evidence, due to the epidemiological design of these studies (which are predominantly case-control studies), they cannot convincingly infer causality between the gut microbiome and esophageal cancer, or the likely direction of any causal effect. Specifically, reverse causation (i.e., where the gut microbial profile is a consequence of esophageal cancer instead of a cause) and confounding (i.e., where the gut microbial profile and esophageal cancer share a common cause but are not inherently causally related), are major limitations of these studies. Furthermore, most existing studies are limited by small sample sizes, reducing their power to detect associations.

Mendelian randomization (MR) is an established study design which uses genetic variants as proxies for a modifiable exposure (e.g., the gut microbiome) to improve causal inference in the relationship between that exposure and an outcome of interest (e.g., esophageal cancer).(19) Firstly, according to Mendel’s laws(19), as genetic variants are randomly assigned at conception, they are not typically associated with common confounders of the exposure and outcome.(20) Secondly, as genetic variants are fixed at conception, they precede disease onset; therefore, using them as proxies for an exposure reduces the risk of reverse causation. In addition, two-sample MR facilitates the use of summary statistics, typically from large genome-wide association studies (GWASs), often increasing statistical power.

There are an increasing number of studies to date using MR to assess causal relationships between the gut microbiome and different health outcomes, including esophageal cancer.(21) However, given the complexity of the relationship between host genetics and the gut microbiome, robust examination of violations of MR assumptions and careful critique of these results is necessary. In this study, we performed a bidirectional, two-sample MR analysis to estimate the direction and magnitude of any causal relationship between the gut microbiome and esophageal cancer (specifically, EA, BE and a combination of the two), appropriately applying a series of sensitivity analyses to test the robustness of observed effects.

## METHODS

### Study design

Two-sample bidirectional MR was used to investigate the causal relationship between the gut microbiome and esophageal cancer. Single nucleotide polymorphisms (SNPs) associated with both the exposure and outcome of interest were obtained from summary data of two independent GWASs of European populations.(9,22) This data was used to analyse the causal effect of 14 microbial traits on EA, BE and both EA and BE as a combined outcome (BE/EA) – i.e., referred to as the “first direction” throughout this study. Then, this data was also used to analyse the causal effect of EA, BE and BE/EA on those same 14 microbial traits – i.e., referred to as the “reverse direction” throughout this study. This study followed the Strengthening the Reporting of Observational Studies in Epidemiology using Mendelian Randomization (STROBE-MR) guidelines.(23)

### Gut microbiome GWAS data and instrument selection

The microbial summary-level data were obtained from a large microbiome GWAS (mGWAS) meta-analysis conducted in three independent European cohorts, including the Flemish Gut Flora Project (FGFP; n=2,223) and two German cohorts (FoCus (n=950) and PopGen (n=717) (data available here: https://data.bris.ac.uk/data/dataset/22bqn399f9i432q56gt3wfhzlc). (22) This mGWAS assessed the association between genetic variants and two types of microbial phenotype characterised by either the presence vs. absence (P/A) or relative abundance (AB) of a microbial trait. Full details on the methodology used by the study are described elsewhere.(22,24)

Briefly, microbiome phylogenetic profiling was characterised with 16S ribosomal RNA gene sequencing of participants’ faecal samples. Following quality control (QC), 92 taxa were available for the mGWAS analysis. If a taxon was absent in the population in more than 5% of cases, a hurdle binary (HB) analysis was performed, consisting of two steps. Firstly, all non-zero counts were converted to 1’s and the mGWAS assessed associations of SNPs with P/A of the microbial trait. Secondly, all the absent cases were removed, and the remaining data were treated as continuous variables and rank normal transformed (RNT) before the mGWAS was performed, investigating associations of SNPs with relative abundance of the microbial trait. Thes steps provided a total of 159 microbial traits available for analysis in the FGFP discovery cohort. All SNPs that reached the pre-determined p-value threshold (P<1x10^-05^) were then included in the meta-analysis including the FoCus and PopGen studies. Six traits were not present in these studies so were not analysed. The meta-analysis was performed using the inverse variance fixed effects method and SNPs were considered “meta-supported” if they reached a smaller p-value of association in the meta-analysis than in the FGFP cohort alone. A Bonferroni correction was also applied, accounting for the number of microbial traits analysed in the mGWAS, where the genome-wide meta-analysis p-value threshold was set at 2.5×10^−08^ and the study-wide p-value threshold was defined as 1.57×10^−10^. Two SNPs reached the study-wide p-value threshold, while a further 11 SNPs reached the genome-wide meta-analysis threshold. For the current MR analyses, one additional SNP was also selected due to the consistently reported association with the bacteria in the *Bifidobacterium* genus in the literature. (25–29)

This gave a total of 14 SNPs, each representing one independent microbial trait, that were used as genetic instruments (or proxies) within MR analyses in the first direction (**Supplementary Table 1**) – i.e., estimating the effect of the gut microbiome on esophageal cancer outcomes. Effect estimates from the mGWAS represent an increase in the log-transformed odds ratio (OR) for P/A microbial traits and the standard deviation (SD) change for rank normalised AB microbial traits with each effect allele carried.

### Esophageal cancer GWAS data and instrument selection

Summary-level data for esophageal cancer were obtained from the largest esophageal cancer GWAS meta-analysis to date.(9) This meta-analysis used six cohorts, comprising 16,790 patients (11,208 BE cases and 5,582 EA cases) and 32,476 ethnically matched controls (9), all of European origin. The following cohorts were used in the meta-analysis: Cambridge, Oxford, Bonn V1, Barrett’s and Esophageal Adenocarcinoma Consortium (BEACON), Bonn V2 and UK Biobank (details of studies available in **Supplementary Table 2**). All patients had a BE or EA histopathological confirmed diagnosis.

Detailed description of the genotyping, imputation, QC and analyses has been previously published.(9,30,31) Ultimately, 18 and seven independent SNPs were associated with BE and EA, respectively, identified at the conventional genome-wide p-value threshold of 5x10^-08^, two of which (one associated with each phenotype) were novel. When combining BE and EA into one phenotype, 24 independent SNPs reached the genome-wide p-value threshold (5x10^-08^), of which nine were novel. These SNPs were used as genetic instruments for the MR analysis in the reverse direction (**Supplementary Tables 3-5**) – i.e., estimating the effect of the esophageal cancer phenotypes on the gut microbiome. Effect estimates from the GWAS represent an increase in the log-transformed OR for each esophageal cancer phenotype with each effect allele carried.

### Statistical analyses

Two-sample MR analyses were completed using the TwoSample MR package (version 0.6.8) in R(32) in the two directions. Firstly, to investigate the causal effect of 14 microbial traits on the risk of BE, EA and BE/EA combined, and secondly to investigate the causal effect of BE, EA and BE/EA combined on the 14 microbial traits.

In the first direction, summary-level data for the 14 SNPs associated with the 14 microbial traits were extracted from the mGWAS and esophageal cancer GWAS meta-analysis. In the reverse direction, summary-level data for the 18, seven and 24 SNPs used as genetic instruments for BE, EA and BE/EA, respectively, were extracted from the GWAS analyses conducted in the FGFP cohort (as only full summary statistics were available for this cohort due to the mGWAS design) and esophageal cancer GWAS meta-analysis. Full details of the methods used here have been previously published.(33) In both directions, if a SNP associated with any exposure was not present in the outcome GWAS summary-level data, a proxy SNP that was in high correlation with the original SNP (r^2^ >0.8) was used, if available.

Prior to MR analyses, SNPs across the exposure and outcome data were harmonised using the “harmonise_data” function on the TwoSampleMR R package,(32) ensuring that the effects of the SNP on the exposure and outcome corresponded to the same allele. For “palindromic” or “ambiguous” SNPs (i.e., those that have effect/other alleles of A/T, respectively, or C/G, respectively) the effect allele was inferred if the SNP has a minor allele frequency (MAF) under 0.42; otherwise, if the MAF>0.42, the SNP was removed from analyses.

In the first direction, as each microbial trait was represented by one SNP, the Wald ratio (WR) method was used to estimate causal effects. This was calculated by dividing the SNP-outcome effect by the SNP-exposure effect.(34) In the reverse direction, as BE, EA and BE/EA all had multiple genetic instruments, the inverse weighted variance (IVW) method was used to estimate causal effects, which takes the mean of the WR estimates that are weighted based on the inverse variance of the association between the instrument and outcome.(35)

As it is likely that the microbial traits analysed in this study were correlated and that there is a high level of genetic correlation between BE, EA and BE/EA, a multiple testing correction was not performed. When considering all performed analyses, the effect estimates coupled with the precision of these estimates (i.e., p-values) were used as a continuous indicator of evidence strength, where those results with the largest effect estimates and smallest p-values were discussed.

### Assumptions of MR analyses

There are three core assumptions of MR analyses (**Figure 1**). Firstly, the genetic variants being used as instruments are robustly associated with the exposure. Secondly, there are no confounders of these genetic instruments and the outcome. Thirdly, the genetic instruments only affect the outcome of interest through the exposure.(36–38) The use of a stringent p-value threshold when selecting genetic instruments increases confidence that the first assumption is met. Additionally, the use of independent SNPs as instruments and samples which represent the same underlying population (i.e., of European ancestry) is likely to minimise the risk of instruments violating the second MR assumption through linkage disequilibrium (LD) or population stratification.

**Figure 1.**
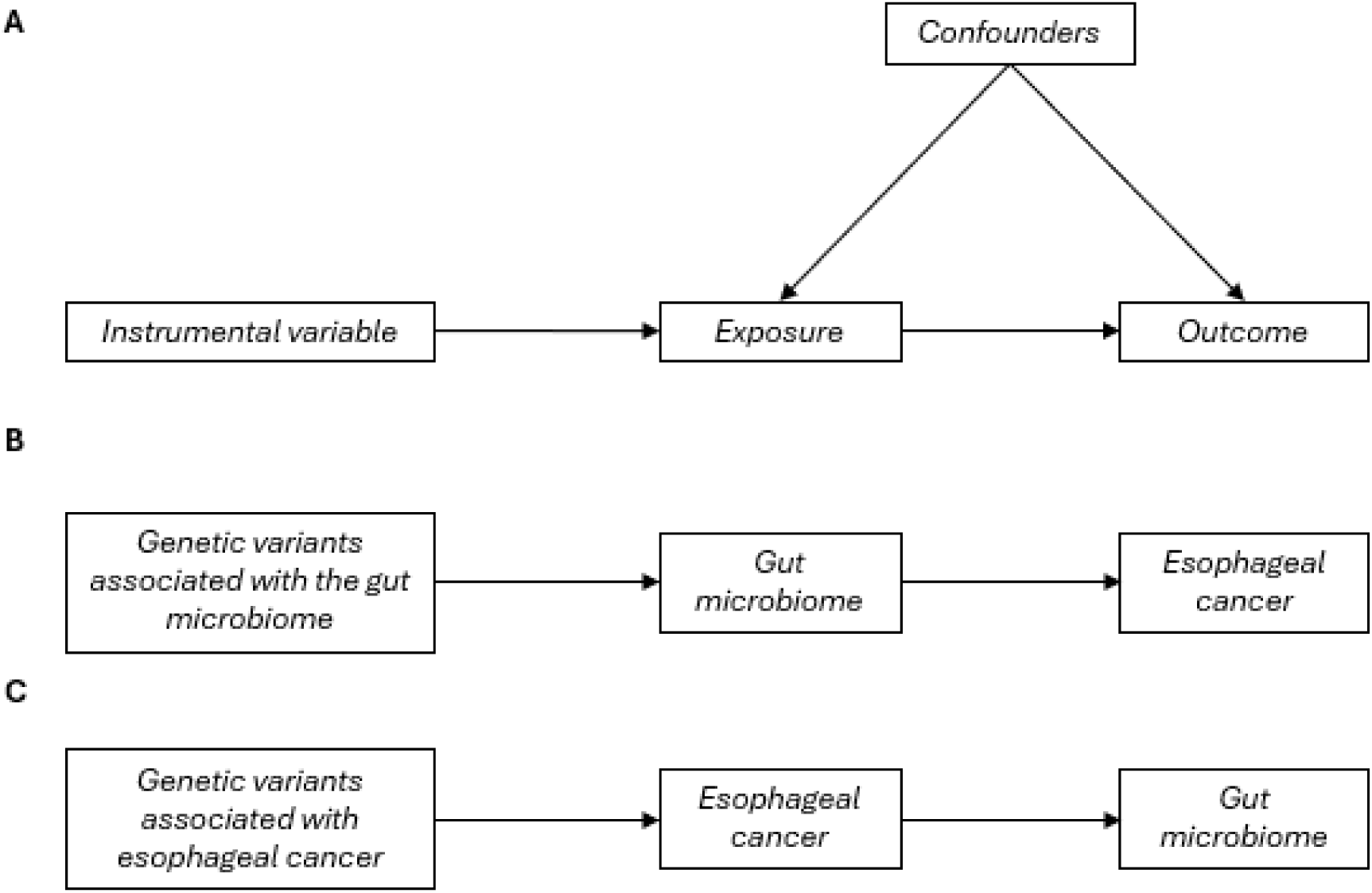
MR framework applied to assess the causal relationship between the gut microbiome on esophageal cancer. MR = Mendelian randomization. Directed acyclic graph (DAG) of the Mendelian randomization (MR) study design. (A) Conceptual MR causal inference framework using genetic instrumental variables (i.e., single nucleotide polymorphisms (SNPs)) robustly associated with the exposure (“relevance” assumption), where there are no confounders of the instrument and outcome (“independence” assumption) and the genetic instruments only affect the outcome through the exposure (“exclusion-restriction” or “no horizontal pleiotropy” assumption). (B) DAG of the MR analysis used to investigate the causal effect of the gut microbiome on esophageal cancer (first direction in this study) and (C) DAG of the MR analysis to investigate the causal effect of esophageal cancer on the gut microbiome (reverse direction in this study).

In both directions, where there was evidence of an exposure having a causal effect on an outcome, we conducted several sensitivity analyses to test the validity of primarily the third MR assumption. In the first direction, these analyses included colocalization, a manual exploration of horizontal pleiotropy and repeating MR analyses with a more lenient p-value for instrument selection (as previously described(33,39)) to enable to use of pleiotropy-robust methods where multiple genetic variants are present. Given that there were several SNPs associated with the esophageal cancer phenotypes for analyses in the reverse direction, we were able to perform these pleiotropy-robust methods including the weighted median(40), the weighted mode (41) and MR-Egger regression(42). Cochran’s Q statistic(43) was used to assess heterogeneity amongst causal estimates across SNPs. Additionally, scatter plots, forest plots, leave-one-out analysis and funnel plots(34) were used to visualise whether directional horizontal pleiotropy was biasing results. Finally, the Steiger(44) test was used to assess the likely direction of causality.

## RESULTS

### First direction

Of the 14 SNPs associated with the 14 microbial traits, seven, eight and seven were available in the BE, EA and BE/EA summary statistics, respectively, and no suitable proxy SNPs could be found.(11) Two-sample MR analyses found little evidence that any of these microbial traits effected EA, BE or BE/EA (**Table 1**, **Figure 2**). Hence, no sensitivity analyses were conducted in the first direction.

**Figure 2.**
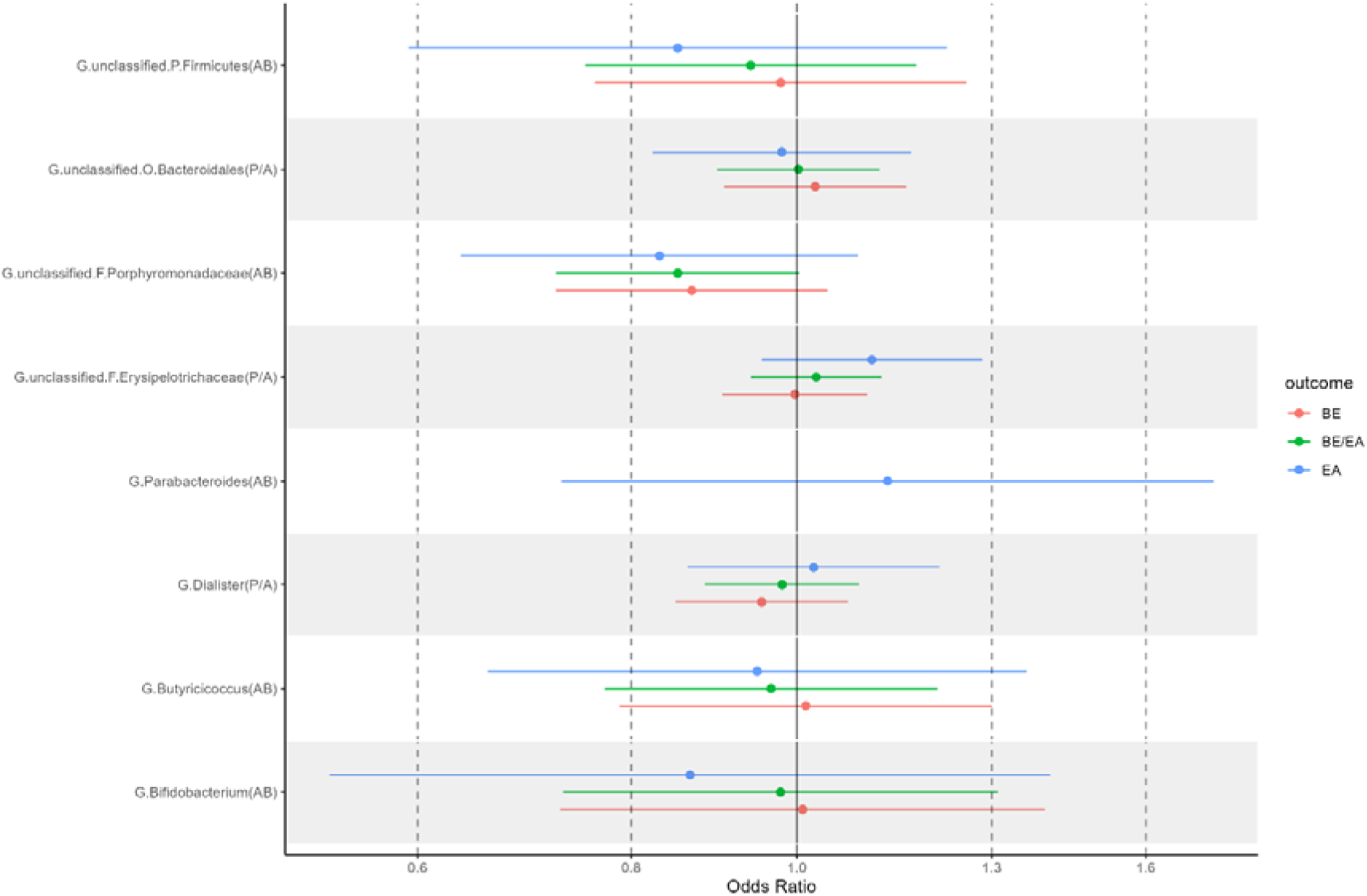
Two-sample MR estimates of the effect of the gut microbiome on esophageal cancer. MR = Mendelian randomization, BE/EA = Barrett’s esophagus and esophageal adenocarcinoma combined, EA = esophageal adenocarcinoma, BE = Barrett’s esophagus, AB = relative abundance, P/A = presence vs. absence. Letters preceding the microbial trait name represent the following: C = class, F = family, G = genus, O = order, P = phylum. Microbial traits that are specified as “unclassified” are traits that could not be classified at the genus level, and therefore have been moved into an unclassified group within the classification above. MR estimates represent the OR for BE/EA, EA and BE risk and 95% CI per SD unit change in relative abundance (AB) for continuous microbial traits or the presence vs. absence (P/A) of each binary microbial trait. Only one effect estimate could be calculated for G.Parabacteroides (AB) as neither the associated SNP nor suitable proxy SNPs could be found in the BE and BE/EA GWAS summary statistics. Effect estimates were unable to be calculated for six microbial traits as neither the associated SNPs nor suitable proxy SNPs could be found in the EA,BE or BE/EA GWAS summary statistics.

**Table 1.** Two-sample MR estimates of the effect of the gut microbiome on esophageal cancer.

| Microbial Trait | MR estimate using the Wald ratio (95% CI); p-value |  |  |
| --- | --- | --- | --- |
|  | BE/EA | EA | BE |
| G.Unclassified.F.Erysipelotrichaceae (P/A) | 1.03 (0.94, 1.12);<br>0.56 | 1.11 (0.95, 1.28); 0.18 | 1.00 (0.90, 1.10);<br>0.96 |
| G.Unclassified.O.Bacteroidales (P/A) | 1.00 (0.90, 1.12);<br>0.97 | 0.98 (0.82, 1.17);<br>0.82 | 1.03 (0.91, 1.16);<br>0.69 |
| G.Dialister (P/A) | 0.98 (0.88, 1.09);<br>0.71 | 1.02 (0.86, 1.21);0.80 | 0.95 (0.85, 1.07);<br>0.42 |
| G.Bifidobacterium (AB) | 0.98 (0.73, 1.31);<br>0.88 | 0.87 (0.53, 1.41);<br>0.56 | 1.01 (0.73, 1.40);<br>0.96 |
| G.Butyricoccus (AB) | 0.97 (0.77, 1.21);<br>0.76 | 0.95 (0.66, 1.36);<br>0.77 | 1.01 (0.79, 1.30);<br>0.92 |
| G.Unclassified.P.Firmicutes (AB) | 0.94 (0.75, 1.17);<br>0.59 | 0.85 (0.60, 1.22);<br>0.39 | 0.98 (0.76, 1.26);<br>0.87 |
| G.Unclassified.F.Porphyromonadaceae (AB) | 0.85 (0.72, 1.00);<br>0.05 | 0.83 (0.64, 1.09);<br>0.18 | 0.87 (0.72, 1.04);<br>0.13 |
| G.Ruminococcus (P/A) | n/a | n/a | n/a |
| G.Coproccoccus (P/A) | n/a | n/a | n/a |
| F.Sutterellaceae (P/A) | n/a | n/a | n/a |
| G.Parabacteroides (AB) | n/a | 1.13 (0.73, 1.75);<br>0.59 | n/a |
| C.Gammaproteobacteria (AB) | n/a | n/a | n/a |
| G.Unclassified.P.Firmicutes (P/A) | n/a | n/a | n/a |
| G.Veillonella (P/A) | n/a | n/a | n/a |

*MR = Mendelian randomization, BE/EA = Barrett's esophagus and esophageal adenocarcinoma, EA =* *esophageal adenocarcinoma, BE = Barrett's esophagus, AB = relative abundance, P/A = presence vs. absence,* *SD = standard deviation, CI = confidence interval. Letters preceding the microbial trait name represent the* *following: C = class, F = family, G = genus, O = order, P = phylum. Microbial traits that are specified as* *"unclassified" are traits that could not be classified at the genus level, and therefore have been moved into an* *unclassified group within the classification above. The effect estimate is the odds ratio (OR) for BE/EA, EA and* *BE risk with either the approximately doubling of the genetic liability to presence vs. absence (P/A) of each binary* *microbial trait or the standard deviation (SD) change in relative abundance (AB) of each continuous microbial* *trait. Entries of "n/a" indicate that the genetic variant associated with that gut microbial trait was not available* *within the GWAS summary statistics and it was not possible to identify a proxy SNP. Therefore, MR analyses* *were not possible for these microbial traits and esophageal cancer outcomes.*

### Reverse direction

In the reverse direction, two-sample MR analyses found evidence that an approximate doubling of the genetic liability to EA increased the odds of presence (vs. absence) of an unclassified group of genus-level taxa within the *Firmicutes* phylum (OR: 1.66; 95% CI: 1.02, 2.70) and decreased the relative abundance of bacteria within the *Butyricicoccus* genus by 0.23 SDs (95% CI: 0.07, 0.40) (**Table 2**, **Figures 3A** and **3B**). There was little evidence that EA affected any of the other microbial traits nor that either BE or BE/EA affected any of the microbial traits analysed. Therefore, sensitivity analyses focused on these two results found in main analyses.

**Figure 3.**
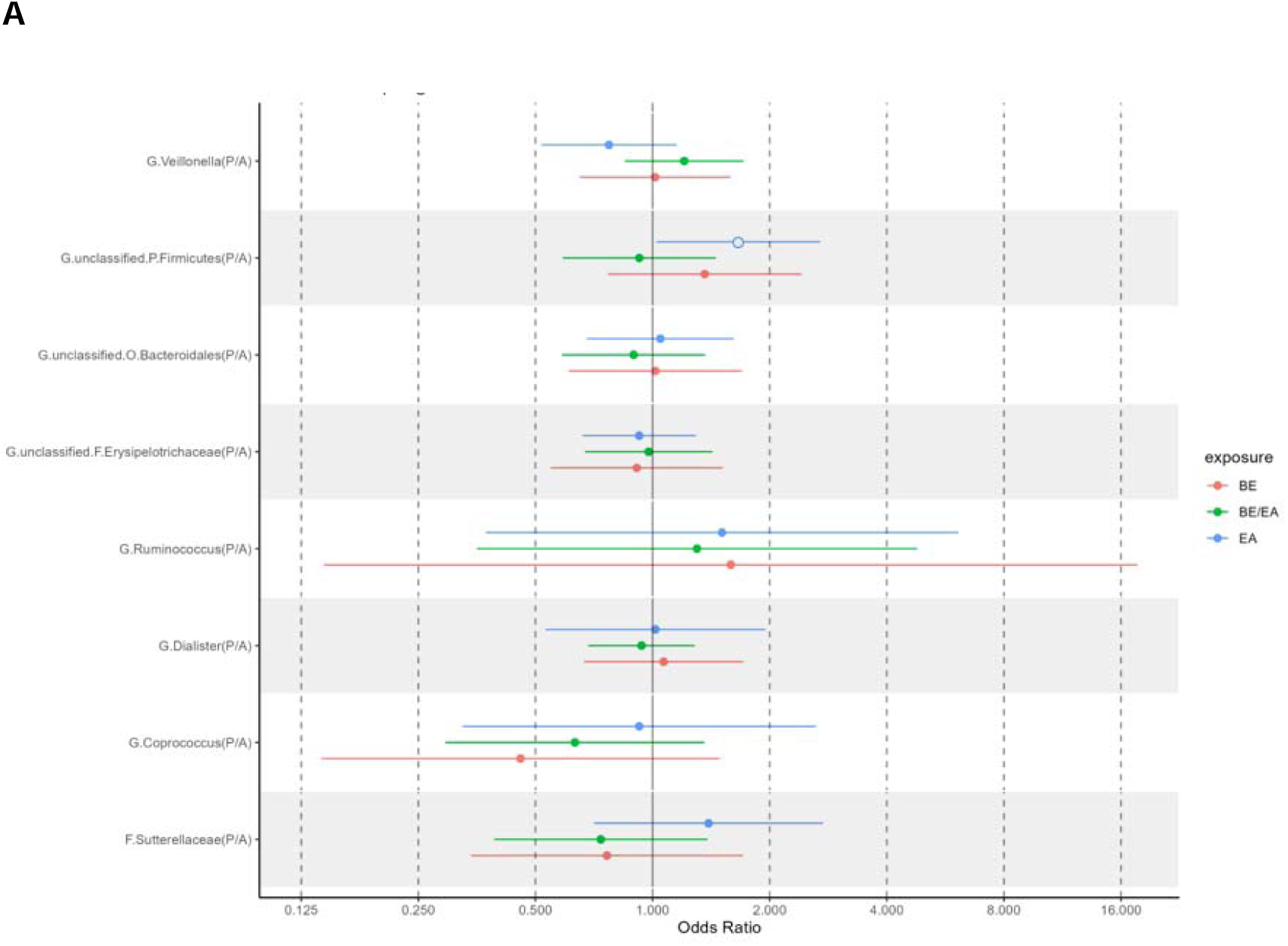
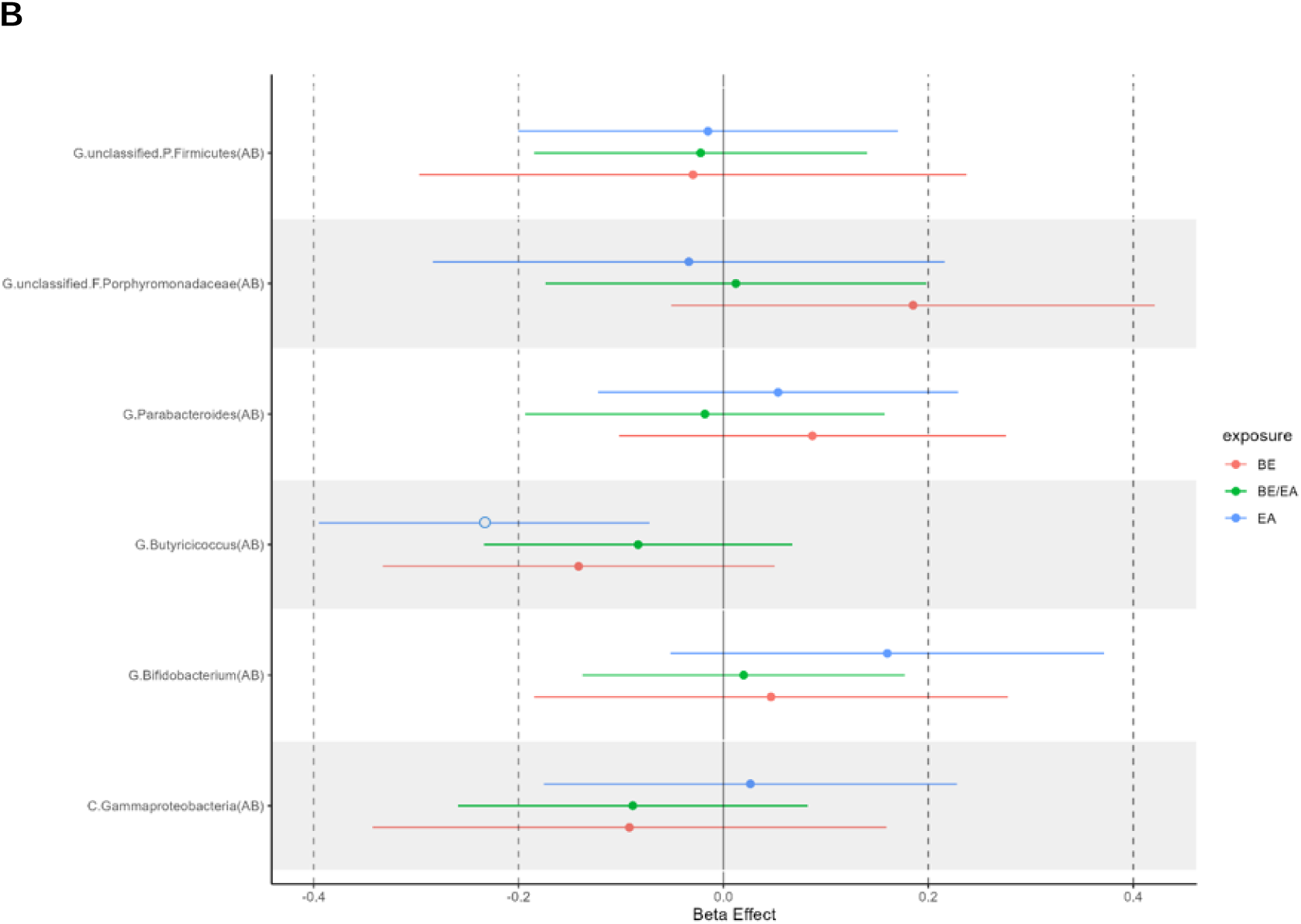
Two-sample MR estimates of the effect of esophageal cancer on the gut microbiome. **A** MR = Mendelian randomization, BE/EA = Barrett’s esophagus and esophageal adenocarcinoma, EA = esophageal adenocarcinoma, BE = Barrett’s esophagus, P/A = presence vs. absence. MR estimate represents the odds ratio (OR) (and 95% confidence interval (CI)) of presence vs. absence of the binary microbial trait with an approximate doubling of the genetic liability to presence vs. absence of each esophageal cancer trait. MR estimates where the 95% CI did not cross the null are indicated by a non-filled circle. **B** MR = Mendelian randomization, BE/EA = Barrett’s esophagus and esophageal adenocarcinoma, EA = esophageal adenocarcinoma, BE = Barrett’s esophagus, AB = relative abundance. MR estimate represents the standard deviation (SD) change in relative abundance (and 95% confidence interval (CI)) of the continuous microbial trait with an approximately doubling of the genetic liability to presence vs. absence of each esophageal cancer trait. MR estimates where the 95% CI did not cross the null are indicated by a non-filled circle.

**Table 2.** Two-sample MR estimates of the effect of esophageal cancer on the gut microbiome.

| Microbial Trait (outcome) | MR estimate (95% CI); p-value |  |  |
| --- | --- | --- | --- |
|  | BE/EA | EA | BE |
| G.Unclassified.F.Erysipelotrichaceae (P/A) | 0.98 (0.67, 1.42); 0.90 | 0.92 (0.66, 1.29); 0.64 | 0.91 (0.55, 1.51); 0.72 |
| G.Unclassified.O.Bacteroidales (P/A) | 0.89 (0.59, 1.37); 0.60 | 0.97 (0.75, 1.24); 0.79 | 1.02 (0.61, 1.70); 0.95 |
| G.Dialister (P/A) | 0.94 (0.68, 1.28); 0.68 | 1.02 (0.53, 1.95); 0.96 | 1.07 (0.67, 1.71); 0.78 |
| G.Bifidobacterium (AB) | 0.02 (-0.14, 0.18); 0.81 | 0.16 (-0.05, 0.37); 0.14 | 0.05 (-0.19, 0.28); 0.69 |
| G.Butyricicoccus (AB) | -0.08 (-0.23, 0.07); 0.28 | **-0.23 (-0.40, -0.07); 0.005** | -0.14 (-0.33, 0.05); 0.15 |
| G.Unclassified.P.Firmicutes (AB) | -0.02 (-0.18, 0.14); 0.79 | -0.02 (-0.20, 0.17); 0.87 | -0.03 (-0.50, 0.53); 0.95 |
| G.Unclassified.F.Porphyromonadaceae (AB) | 0.01 (-0.17, 0.19); 0.90 | -0.03 (-0.28, 0.22); 0.79 | 0.19 (-0.05, 0.42); 0.12 |
| G.Ruminococcus (P/A) | 1.30 (0.35, 4.78); 0.69 | 1.51 (0.37, 6.12); 0.56 | 1.59 (0.14, 17.65); 0.71 |
| G.Coproccoccus (P/A) | 0.63 (0.29, 1.36); 0.24 | 0.92 (0.32, 2.63); 0.88 | 0.46 (0.14, 1.49); 0.19 |
| F.Sutterellaceae (P/A) | 0.74 (0.39, 1.38); 0.34 | 1.39 (0.71, 2.74); 0.34 | 0.76 (0.34, 1.70); 0.51 |
| G.Parabacteroides (AB) | -0.02 (-0.19, 0.16); 0.84 | 0.05 (-0.12, 0.23); 0.55 | 0.09 (-0.10, 0.28); 0.37 |
| C.Gammaproteobacteria (AB) | -0.09 (-0.26, 0.08); 0.31 | 0.03 (-0.18, 0.23); 0.80 | -0.09 (-0.34, 0.16); 0.47 |
| G.Unclassified.P.Firmicutes (P/A) | 0.92 (0.59, 1.45); 0.73 | **1.66 (1.02, 2.70); 0.04** | 1.36 (0.77, 2.42); 0.29 |
| G.Veillonella (P/A) | 1.21 (0.85, 1.71); 0.30 | 0.77 (0.52, 1.15); 0.21 | 1.02 (0.65, 1.58); 0.95 |

MR = Mendelian randomization, BE/EA = Barrett's esophagus and esophageal adenocarcinoma, EA = esophageal adenocarcinoma, BE = Barrett's esophagus, AB = relative abundance, P/A = presence vs. absence, SD = standard deviation, IVW = inverse variance weighted, CI = confidence interval, OR = odds ratio. Letters preceding the microbial trait name represent the following: C = class, F = family, G = genus, O = order, P = phylum. Microbial traits that are specified as "unclassified" are traits that could not be classified at the genus level, and therefore have been moved into an unclassified group within the classification above. For presence vs. absence (P/A) microbial traits, the MR estimate represents the odds ratio (OR) of presence vs. absence of the binary microbial trait with an approximate doubling in the genetic liability to presence vs. absence of each esophageal cancer trait. For relative abundance (AB) microbial traits, the MR estimate represents the standard deviation (SD) change in relative abundance of the continuous microbial trait with an approximate doubling of the genetic liability to presence vs. absence of each esophageal cancer trait. MR effect estimates where the confidence interval (CI) did not cross the null are indicated with bold text.

### Sensitivity analyses

Estimates of the causal effects of EA on the unclassified group of bacteria within the *Firmicutes* phylum and on the relative abundance of bacteria within the *Butyricicoccus* genus using the weighted median, weighted mode and MR-Egger estimates were all in the same direction and consistent with the IVW estimates (**Supplementary Table 6**, **Supplementary** Figures 1A-B, **Supplementary** Figures 2A-B), though with expectedly large confidence intervals around the MR-Egger estimate. The Cochran’s Q statistics indicated that there was little evidence for heterogeneity amongst instrument-specific causal effect estimates (P=0.71 for the *Firmicutes* trait and P=0.47 for the *Butyricicoccus* trait; **Supplementary Table 6**).

However, leave-one-out analyses showed that the originally observed causal effect of EA on this unclassified group of bacteria within the *Firmicutes* phylum was likely driven in part by each individual SNP, as removing almost all SNPs in turn attenuated the effect estimates to the null (apart from rs9880983 and rs10431648, where the original SNP-level effect estimates of which overlapped the null in the main analysis) (**Supplementary** Figure 1C). In the analyses of the relative abundance of *Butyricicoccus* trait, leave-one-out analysis showed that removing almost each SNP in turn did not have a large impact on the observed effect estimate; however, removing one SNP (rs376563, which had the largest SNP-level WR estimate in the main analyses) did attenuate the effect towards the null, suggesting that the initially observed causal effect may have been driven by this SNP individually (**Supplementary** Figure 2C).

Whilst there was limited statistical evidence for an effect of unbalanced pleiotropy (MR-Egger intercept = -0.19; P=0.64 for the *Firmicutes* trait and -0.01; P=0.94 for the *Butyricicoccus* trait; **Supplementary Table 6**), this was likely driven by the small number of SNPs in these analyses (n=7), which was emphasized further with the scarcity and asymmetry of the funnel plots (**Supplementary** Figures 1D and 2D). Lastly, the Steiger test suggested that both effect estimates were in the correct hypothesized causal direction (P=0.17 for the *Firmicutes* trait and P=0.31 for the *Butyricicoccus* trait; **Supplementary Table 6**) – i.e., that the SNPs explained more variation in esophageal cancer than in these gut microbial traits), meaning that there was little evidence that reverse causality was biasing the IVW causal effect estimate.

## DISCUSSION

In this study, we used two-sample bidirectional MR analyses to explore the causal relationship between the gut microbiome and esophageal cancer. Firstly, our MR analyses provided little evidence of a causal effect of any microbial trait on EA, BE and EA/BE outcomes. Secondly, whilst, in the reverse direction, MR analyses provided initial evidence that a higher genetic liability to EA increased the odds of the presence of an unclassified group of bacteria in the *Firmicutes* phylum and decreased the relative abundance of bacteria within the *Butyricicoccus* genus, sensitivity analyses indicated that these results were not robust when assessing violations of MR assumptions. Specifically, whilst there was little evidence of statistical heterogeneity between SNP-level estimates within these analyses, of unbalanced horizonal pleiotropy or of reverse causality based on the Cochran’s Q test, MR-Egger intercept and Steiger test, respectively, there were firstly few SNPs in these analyses, each of which seemed to drive the observed effect individually (which was clear upon removal of each SNP in turn within the leave-one-out analysis) to varying extents. Therefore, the initial observed effects of EA on these microbial traits are likely to be complex and not reflective of causality and thus needs to be taken with caution.

There are a small number of observational studies exploring the gut microbiome and esophageal cancer, often with conflicting results.(17,45–48) For example, some studies have reported a higher prevalence of microbiota in the *Firmicutes* phylum amongst patients with esophageal cancer, aligning with our initial findings in the reverse direction. (17,46–48) Additionally, a higher abundance of bacteria in the *Butyricicoccus* genus amongst patients with esophageal cancer has also been reported.(17) The findings from these case-control studies in isolation could be interpreted as a relationship between these microbial traits and the risk of esophageal cancer; however, these studies are unable to discern direction or reliability of any possible causal effect.

A recent similar two-sample MR study, published by Li *et al*. (21), reported evidence for a causal effect of five microbial traits (*Veillonella, Coprobacter, Prevotella9* and *Turicibacter* genus-level taxa and a *Eubacterium oxidoreducens* group of bacteria) on esophageal cancer and two microbial traits (*Flavonifractor* and *Actinomyces* genus-level taxa) on EA, inconsistent with our findings. There are several reasons why these findings were not replicated in our study. Firstly, both our mGWAS and esophageal cancer GWAS contained populations of European ancestries, whereas Li *et al.* used a different mGWAS study, which contained participants of multiple ancestries; thus, increasing the risk of their instruments being confounded by population stratification.(25) Secondly, we used a genome-wide p-value threshold defined by the mGWAS (P < 2.5x10^-08^) (22) to select genetic instruments associated with 14 microbial traits, whereas Li *et al.*(21) used a more lenient threshold (P < 1x10^-05^) to select instruments that were associated with 196 microbial traits. Subsequently, the microbial traits on which Li *et al.* focused were not analysed in this current paper. Additionally, as opposed to other gut microbial MR analyses including Li *et al.*, the genetic instruments used to represent microbial traits in the current study had been consistently associated across multiple cohorts.(22)

Whilst unfortunately increasingly commonplace in the application of MR with microbiome phenotypes, the use of a lenient p-value threshold to select genetic variants associated with microbial traits (where these relationships are often not replicated) increases the risk of bias through weak instrumentation, horizontal pleiotropy and genetic confounding due to violations of core MR assumptions and thus is not in adherence with the STROBE-MR framework for improving causal inference. Finally, Li *et al.* did not conduct reverse MR analyses or comprehensive sensitivity analyses (e.g., the Steiger test), which limited their ability to assess how likely reverse causality may explain observed effects (e.g., that the SNPs associated with a microbial trait actually influence esophageal cancer, which, in turn, influences the microbial trait, which could be formally tested through reverse MR analyses).

Importantly, comparing our study with these previously published papers has its limitations, given the demographic difference in cohorts and lack of distinction between histological subtypes of esophageal cancer, which is important given the rise of EA, specifically in the Western world.(8) As the prognosis of esophageal cancer remains poor, the most striking factor being the stage of the cancer at diagnosis (4), earlier detection of esophageal cancer, particularly using non-invasive and easily accessible testing, will be key in reducing the disease burden. Indeed, gut microbial markers for other diseases are becoming more widely researched (49), including those that have identified potential microbial biomarkers for hepatocellular carcinoma.(50) Therefore, in addition to assessing the potential causal role of the gut microbiome on health outcomes (for which comprehensive and reliable MR analyses are currently unable to provide convincing evidence given the limited number of associated SNPs available from large mGWASs), further analyses focusing on the impact of cancer on the gut microbiome may prove more relevant for the identification of clinically meaningful biomarkers for the diagnosis and progression of cancer.

The strengths of this study align to the adherence to the three core assumptions underpinning any MR study (**Figure 1**). The use of stringent p-value thresholds to select genetic instruments increases our confidence that the first “relevance” assumption is met and we used multiple sensitivity analyses to explore the validity of the second and third assumptions. Additionally, performing bidirectional analysis and the Steiger tests allowed us to assess the likelihood that reverse causality was biasing our results and quantify the likely direction of observed effects. Lastly, we used the largest esophageal cancer GWAS to date, increasing our power to detect modest effects, and considered different histological subtypes of esophageal cancer. (9)

However, this study has several limitations relating to the current utility of MR in understanding the role played by the gut microbiome in relation to health phenotypes given the currently limited knowledge of host genetic effects on the microbiome and number of SNPs available for such analyses. Firstly, there are many organisms of lower taxonomic class within the characterised microbial traits that could be driving (or indeed masking) observed associations. Any future efforts of harnessing observed effects derived from MR analyses as either a preventative, diagnostic or therapeutic target would require much more precise classifications of the exact species and strains that could be responsible for any observed relationship. Although we did not find any strong evidence for a causal effect of any microbial traits on esophageal cancer, this may because of the modest sample size of the mGWAS used, limiting our power to detect them. This emphasises the need for more informative microbial sampling techniques and harmonized analytical methods used within future larger mGWASs that would enable the identification of lower taxonomical specificity and more SNPs robustly associated with such microbial traits. Lastly, we only used data from individuals of European ancestry and EA histology, and so our findings may only be relevant to this population.

## CONCLUSIONS

In conclusion, our study provided little evidence that gut microbial traits play a causal role in esophageal cancer aetiology and, whilst initial evidence suggested a causal role of EA in increasing the presence of certain bacteria within the *Firmicutes* phylum and decreasing the relative abundance of bacteria within *Butyricicoccus* genus, sensitivity analyses suggested that these results may have been driven by horizontal pleiotropy. Therefore, the initial observed effects of EA on these microbial traits are likely to be complex and not reflective of causality and thus need to be taken with caution. Our study further highlights the need for caution in the application and assessment of MR analyses and, specifically, a requirement for a more direct assessment of the causal direction of these associations (indeed, if there is a causal relationship). Importantly, MR is a single method, with unique strengths and limitations, that cannot prove causality in isolation. Like any epidemiological study design, MR has the most value in providing insight into causality and directionality of relationships when combined with other study designs with orthogonal biases.

## List of abbreviations

(BE): Barrett’s esophagus
(BEACON): Barrett’s and Esophageal Adenocarcinoma Consortium
(EA): esophageal adenocarcinoma
(BE/EA): esophageal adenocarcinoma and Barrett’s esophagus as a combined outcome
(ESCC): esophageal squamous cell carcinoma
(FGFP): Flemish Gut Flora Project
(GERD): gastro-esophageal reflux disease
(GWASs): genome-wide association studies
(HB): hurdle binary
(IVW): inverse weighted variance
(LD): linkage disequilibrium
(MR): Mendelian randomization
(mGWAS): microbiome genome-wide association studies
(MAF): minor allele frequency
(OR): odds ratio
(P/A): presence vs. absence
(QC): quality control
(RNT): rank normal transformed
(AB): relative abundance
(SNPs): single nucleotide polymorphisms
(SD): standard deviation
(STROBE-MR): Strengthening the Reporting of Observational Studies in Epidemiology using Mendelian Randomization
WR: Wald ratio

## DECLARATIONS

### Ethics approval and consent to participate

Informed consent and appropriate ethical approval were obtained for each genome-wide association study (GWAS) used in this study. As only summary-level statistics were used here, no further ethical approval or consent was required.

### Consent for publication

Not applicable.

### Availability of data and materials

GWAS summary-level data for the gut microbiome used in this study were publicly available from the microbiome GWAS (mGWAS) conducted by Hughes *et al*.(22) – also available here: https://data.bris.ac.uk/data/dataset/22bqn399f9i432q56gt3wfhzlc. Summary-level data for esophageal cancer outcomes were obtained by the GWAS conducted by Schröder *et al*. (9), which are available from the corresponding author on reasonable request. As no further datasets were generated in the current study, data sharing is not applicable to this article.

### Competing interests

The authors declare that they have no competing interests.

### Funding

CH was funded by a Cancer Research UK (CRUK) Population Research Postdoctoral Fellowship (RCCPDF\100007), awarded to KHW. KHW is funded by the University of Bristol and is affiliated to the CRUK Integrative Cancer Epidemiology Programme (ICEP; C18281/A29019) and Medical Research Council Integrative Epidemiology Unit (MRC-IEU; MC_UU_00011/1). For the purpose of open access, the author(s) has applied a Creative Commons Attribution (CC BY) licence to any Author Accepted Manuscript version arising from this submission for the purposes of publishing and open access.

### Authors’ contributions

KHW and JC conceived and designed the work and acquired the appropriate data; KHW, JC, CH and GH analysed the data; and all authors interpreted the results and drafted, revised and approved the manuscript.

## Supporting information

STROBE-MR Checklist

## Data Availability

GWAS summary-level data for the gut microbiome used in this study were publicly available from the microbiome GWAS (mGWAS) conducted by Hughes et al.(22) - also available here: https://data.bris.ac.uk/data/dataset/22bqn399f9i432q56gt3wfhzlc. Summary-level data for esophageal cancer outcomes were obtained by the GWAS conducted by Schröder et al. (9), which are available from the corresponding author on reasonable request. As no further datasets were generated in the current study, data sharing is not applicable to this article.

## Acknowledgements

We thank Dr Carlo Maj, Professor Johannes Schumacher and Dr Claire Palles for sharing the GWAS summary-level data for esophageal cancer outcomes for this study.

## Notes

### Competing Interest Statement

The authors have declared no competing interest.

