## Supplementary Figures for "Application of bidirectional Mendelian randomization to assess the relationship between the gut microbiome and esophageal cancer"

**Supplementary Figure 1: MR results for the effect of esophageal adenocarcinoma on G.Unclassified.P.Firmicutes (P/A)**

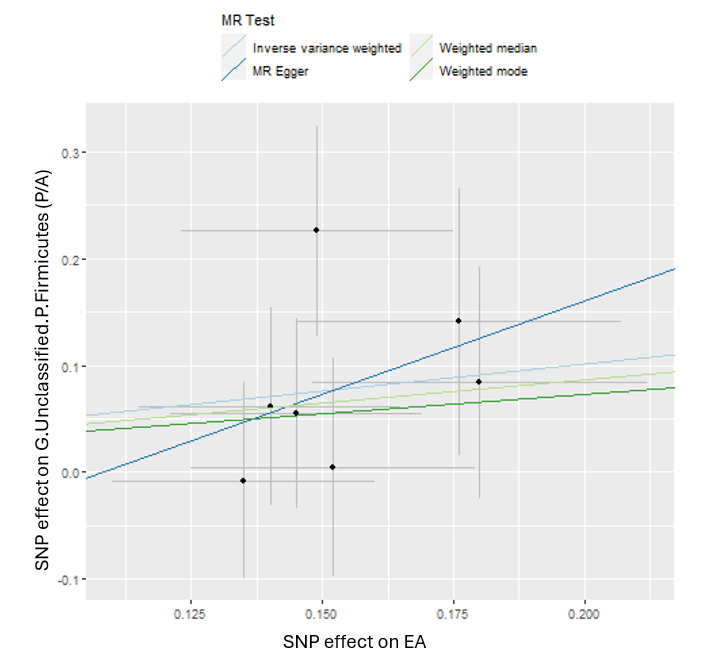

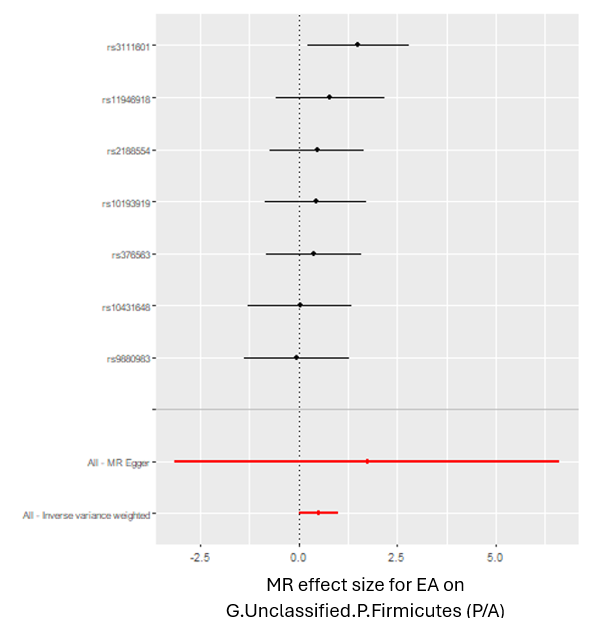
**A	B**

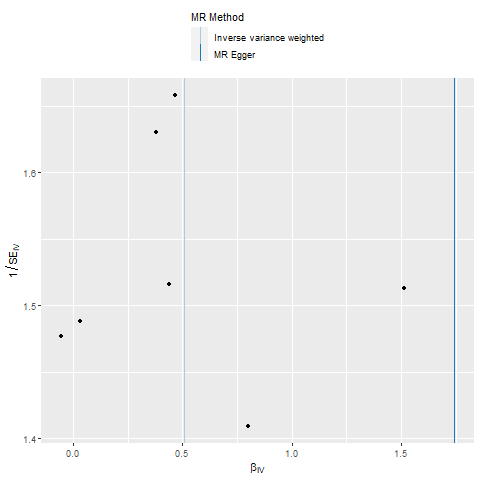
**C	D**

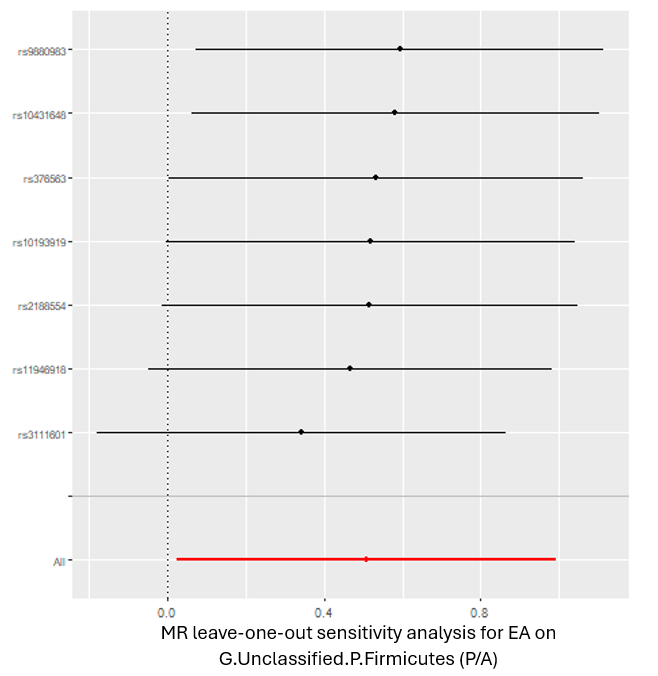

*MR = Mendelian randomization; SNP = single nucleotide polymorphism; P/A = presence vs. absence. These plots show the results of MR analyses to test the causal effect of a higher genetic liability of esophageal adenocarcinoma (EA) on the presence vs. absence (P/A) of unclassified bacteria within the Firmicutes phylum (G.Unclassified.P.Firmicutes). A) Scatter plot comparing four MR methods; B) forest plot comparing the effect of individual SNP-level effect estimates derived using the Wald ratio method and in combination with the inverse variance weighted (IVW) and MR-Egger estimates; C) leave-one-out analysis to check if any one SNP is driving the causal effect; and D) funnel plot comparing the effect estimate and precision of each SNP-level Wald ratio estimate.*

**Supplementary Figure 2: MR results for effect of esophageal adenocarcinoma on G.Butyricicoccus(AB)**

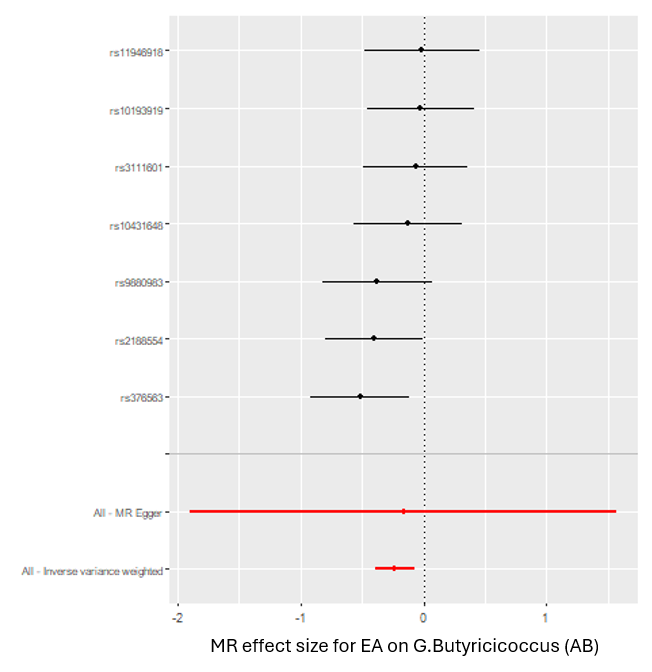

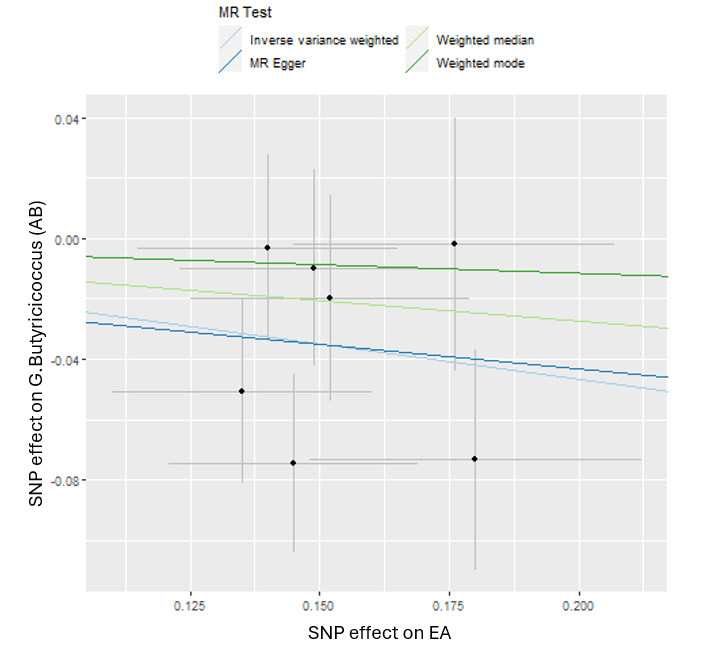
**A	B**

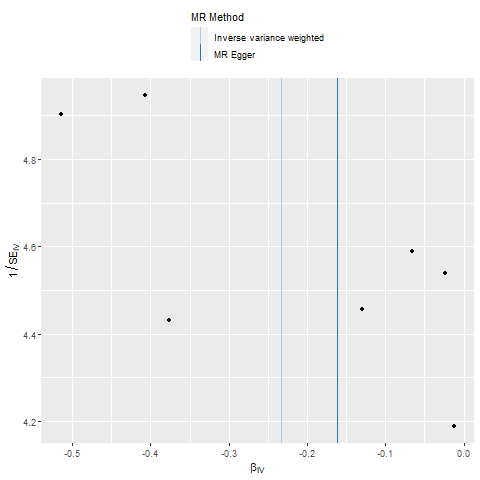

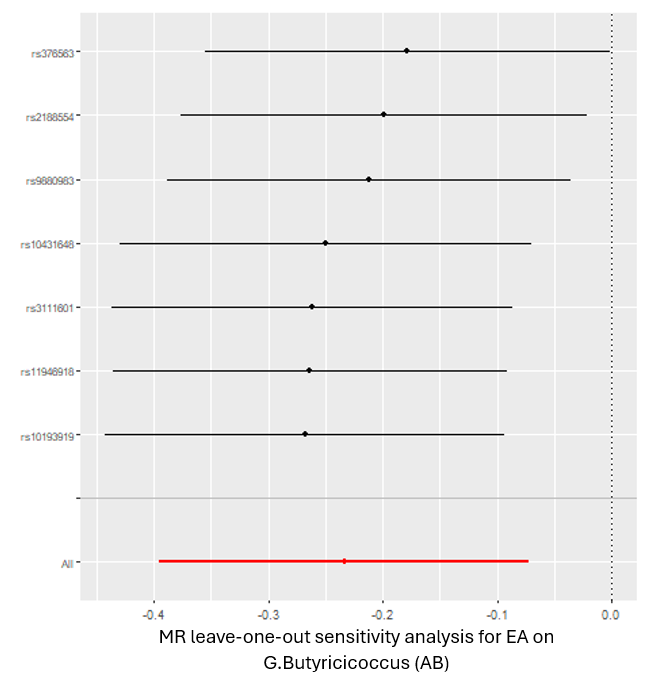
**C	D**

*MR = Mendelian randomization; SNP = single nucleotide polymorphism; P/A = presence vs. absence. These plots show the results of MR analyses to test the causal effect of a higher genetic liability of esophageal adenocarcinoma (EA) on the relative abundance (AB) of bacteria in the Butyricicoccus genus (G.Butyricicoccus). A) Scatter plot comparing four MR methods; B) forest plot comparing the effect of individual SNP-level effect estimates derived using the Wald ratio method and in combination with the inverse variance weighted (IVW) and MR-Egger estimates; C) leave-one-out analysis to check if any one SNP is driving the causal effect; and D) funnel plot comparing the effect estimate and precision of each SNP-level Wald ratio estimate.*
